# Performance and evaluation in computed tomographic colonography screening: protocol for a cluster randomised trial

**DOI:** 10.1101/2020.02.25.20027714

**Authors:** Andrew A Plumb, Anu E Obaro, Paul Bassett, Rachel Baldwin-Cleland, Steve Halligan, David Burling

## Abstract

**Background:** Colorectal cancer (CRC) is a common, important healthcare priority and improving patient outcome relies on early diagnosis. Colonoscopy and computed tomographic colonography (CTC) are commonly-used diagnostic tests. Although colonoscopists are highly regulated and must be accredited, no analogous process exists for CTC. There are currently no universally accepted radiologist performance indicators for CTC, and lack of regulatory oversight may lead to variability in quality and lower neoplasia detection rates. This study aims to determine whether a structured educational training and feedback programme can improve radiologist interpretation accuracy.

**Methods:** NHS England CTC reporting radiologists will be cluster randomised to either an intervention (one-day individualised training and assessment with feedback) or control (assessment with no training or feedback) arm. Each cluster represents radiologists reporting CTC in a single NHS site. Both the intervention and control arm will undertake four CTC assessments at baseline, 1-month (after training; intervention arm or enrolment; control arm), 6- and 12 months to assess their detection of colorectal cancer (CRC) and 6mm+ polyps. The primary outcome will be difference in sensitivity at the 1-month test between arms. Secondary outcomes will include sensitivity at 6 and 12 months and radiologist characteristics associated with improved performance. Multilevel logistic regression will be used to analyse per-polyp and per-case sensitivity. Local ethical and Health Research Authority approval have been obtained.

**Discussion:** Lack of infrastructure to ensure that CTC radiologists can report adequately and lack of consensus regarding appropriate quality metrics may lead to variability in performance. Our provision of a structured education programme with feedback will evaluate the impact of individualised training and identify the factors related to improved radiologist performance in CTC reporting. An improvement in performance could lead to increased neoplasia detection and better patient outcome.

**Registration:** Clinical Trials (ClinicalTrials.gov Identifier: NCT02892721); available from: https://clinicaltrials.gov/ct2/show/NCT02892721. NIHR Clinical Research Network (CPMS ID 32293).

## BACKGROUND

Globally, colorectal cancer (CRC) is the third most commonly diagnosed malignancy and the fourth leading cause of cancer-related deaths [1]. Early stage CRC has excellent cure rates (>95%)[2], whereas late presentation has poor prognosis. Early diagnosis of CRC is therefore crucial, which requires rapid, universal access to accurate, well-tolerated diagnostic tests. For most patients, colonoscopy is used, since it is accurate, widely available and permits confirmatory biopsy of any abnormalities. However, CT colonography (CTC) is an excellent alternative to colonoscopy when colonic investigation is needed [3].

CTC uses standard computed tomography scans to obtain images of the prepared, gas-distended colon and has high sensitivity for the diagnosis of CRC (meta-analysis suggests it is equal to colonoscopy [4]) and large polyps (90% sensitivity for 1cm+ polyps in two separate multicentre prospective cohort studies [5, 6]). A Dutch randomised trial comparing the diagnostic yield of CTC and colonoscopy when screening for CRC found the two tests had equivalent detection rates on a per-invitee basis [7], with CTC outperforming colonoscopy by 3-year follow-up [8]. Notably, the CTC imaging datasets in these pivotal trials were interpreted by trained, experienced radiologists.

CTC has disseminated rapidly across most healthcare systems in the developed world; for example, over 100,000 examinations are conducted each year in the NHS in England alone [9]. Although most radiologists have received basic training in CTC interpretation, there is no routine performance monitoring process to ensure diagnostic accuracy. This situation contrasts with colonoscopy, for which numerous evidence-based metrics including adenoma detection rate (ADR), caecal intubation rate (CIR), withdrawal time (WT) and adverse event rate (AER) are monitored [10]. Moreover, when applied in clinical practice across the English Bowel Cancer Screening Programme (BCSP), detection rates of CTC are generally lower than at colonoscopy; previous research has shown a 50% lower detection rate for both cancers and high-risk polyps [11]. However, hospitals with highly experienced radiologists (>1000 cases), with high throughput (>175 patients / radiologist / annum) and using 3-dimensional interpretation had significantly higher detection rates at CTC, and similar to that at colonoscopy. It is therefore extremely important that we identify methods to improve average CTC interpretive performance.

Although the learning curve at CTC has been previously studied [12], and experienced radiologists are known to be more accurate than novices who have received basic training [13], few studies have addressed whether or not it is possible to use targeted training to improve diagnostic performance among experienced radiologists who are already using CTC in their clinical practice. This is critical, since CTC examinations are conducted across a wide range of hospitals rather than being concentrated in a handful of academic centres [14]. Individual-level randomisation (i.e. at the level of the radiologist) is one possible way to evaluate such training and monitoring; however, radiologists typically work in clinical groups / teams. Training any given radiologist at a particular hospital will inevitably lead to changes in practice, attitude and behaviour among their colleagues. It is therefore not possible to reliably deliver an educational intervention to an individual radiologist and observe its effect in isolation. Therefore, we chose to randomise by cluster to avoid these problems.

This protocol describes the design and methodology of a cluster randomised trial of an individualised training intervention for radiologists to establish (a) whether such training can improve detection of CRC and 6mm+ polyps; (b) the durability of such improvement, if any; (c) factors associated with variability in radiologist diagnostic accuracy; and (d) acceptability of training.

The study is registered (ClinicalTrials.gov Identifier: NCT02892721); available from: https://clinicaltrials.gov/ct2/show/NCT02892721.

## METHODS

### Trial design and setting

This is a parallel group, two-arm, cluster randomised superiority trial with a 1:1 allocation ratio conducted in the UK National Health Service (NHS), aiming to recruit from 50-80 NHS hospital sites. Each eligible NHS CTC reporting site will constitute a cluster; NHS Trusts with multiple physical sites (i.e. multi-hospital Trusts) will be permitted to constitute multiple clusters if their radiologists work separately (to avoid cluster contamination). Individual participants will inevitably be unblinded to arm allocation, since the intervention aims to test individualised training (vs no training), which necessarily requires that participants are aware to which trial arm they have been randomised. Radiologists in both arms will complete four separate evaluations of their interpretative performance; at baseline, one month, 6 months and 12 months. Each of these evaluations will comprise interpretation of 10 different CTC examinations (see Test Datasets section below).

### Eligibility criteria for clusters and individual participants

NHS hospitals currently providing CTC in routine practice (both BCSP and non-BCSP sites) are eligible. Individual participants will be NHS Radiologists reporting for the CTC service, either Consultants or non-consultant grade radiologists (including specialty trainees and clinical fellows) who are within one year of their CCT date (or are post-CCT). Clusters with a mixture of eligible and ineligible individual radiologists will still be permitted to participate, but only eligible radiologists will be included.

We will recruit mainly substantive NHS consultants since this represents the group who interpret the majority of CTC in clinical practice. Furthermore, previous data confirm that this group desire directed training; a survey conducted in 2013 showed that 81% were in favour of accreditation, with the most commonly-suggested strategy being periodic testing with feedback [14]. Although randomisation will occur at cluster level, participant consent will be at the individual level, and participants must complete each test in sequence to be eligible for the subsequent test.

### Test datasets: Composition

Each of the four tests of radiologist interpretative performance will comprise a different set of 10 CTC examinations (“cases”); therefore 40 different cases in total. The cases chosen will be typical of the spectrum of disease encountered in screening practice, with the emphasis on detection of early (i.e. subtle) lesions since these are most likely to be curable. A prevalence of 50-70% abnormal cases per test set (average: 60%) will be used, which is at the upper range of that seen among patients with positive faecal immunochemical testing (FIT) [15]. A relatively high prevalence maximizes statistical power, as it reduces the likelihood of ceiling effects due to all readers identifying most lesions under both conditions (i.e. whether randomised to intervention or control).

All positive cases used for the test datasets will be drawn from a previously collated archive, with colonoscopic and/or histopathological proof. True-positive cases (i.e. those depicting a cancer or polyp that was subsequently proven to be genuine) will be independently scrutinized by four experienced radiologists (average >1000 CTC cases) and visibility, size, segmental location and CT slice numbers of each lesion agreed and documented on a master data sheet, denoting the reference standard for the presence of abnormality.

True-negative CTC datasets will derive from two sources; firstly, those from prior research studies in which both CTC and subsequent colonoscopy were normal; and, secondly, previously-collated cases with no polyp or cancer detected by BOTH of (a) interpretation by two independent expert CT colonographers and (b) minimum of 24 months follow-up with no development of CRC or polyp larger than 5mm. This denotes the reference standard for the presence of normality.

Imaging datasets will be anonymised by a research co-ordinator using a study code and will contain only two scanned positions e.g. supine and prone, or supine and decubitus. Images will then be provided on DVD (via post) for participants to upload to local PACS / CTC workstations. An online platform to allow remote viewing of test cases will be provided by Vitrea Enterprise Suite (Vital Images) if desired.

### Test datasets: Radiologist Interpretation

Radiologists will be asked to identify only intracolonic abnormalities by completing an online Case Report Form (CRF) for each case in each test set. This will capture information regarding the case number, diagnosis; normal (no polyp of 6mm or greater) or abnormal (CRC or polyps larger than 6mm), slice location of any suspected abnormalities, diagnostic confidence (using a 10-point scale), whether or not CAD was used, and proportion of time spent using 2D vs 3D visualisation. To allow for measurement error, radiologists will be asked to document any polyps measured at 5mm or greater, but only polyps with the reference standard size of 6mm or greater will be included for analysis of the study endpoints. Radiologists will also be asked to select the most appropriate management of the case from one of five options:

1. Repeat CT colonography as the current study is suboptimal
2. No additional colonic investigation required
3. CT colonography surveillance within 3 years
4. Refer for consideration of endoscopic evaluation (for characterisation +/- biopsy or polypectomy)
5. Refer for consideration of same day colonoscopy, biopsy and CT staging

The CTC tests for both arms will be marked manually by the central research team using pre-specified agreed parameters (provided below), and the same pre-specified criteria will be universally applied to all participants. An individual polyp or cancer will be regarded as having been detected (i.e. per-polyp unit of analysis) by the interpreting participant if the correct slice number (or numbers) depicting that polyp is stated; and at least two of the following three parameters:

1. Correct classification of the lesion type (polyp – 5mm+, cancer – polypoid, cancer – mass like or other).
2. Correct segment identified (same as the pre-specified segment or the adjacent colonic segment, to allow for inter-observer variability in segment identification).
3. Size measurement within 50% of the true lesion dimension, as defined by the reference standard.

Since individual CTC cases may harbour more than one polyp, we also require a definition of a correctly identified case (i.e. per-case unit of analysis). This will require identification of the “index colorectal lesion”, defined as the neoplastic lesion with the most advanced histology (or, where there are several neoplasms with the same histology, the largest of these); or, for normal cases, no false positive polyps stated (see below).

A false positive lesion is defined as a radiologist participant identifying a polyp or CRC of any size in a colonic segment that is identified as normal as per the reference standard. Diminutive (<6mm) polyps marked by participants that are truly present as per the reference standard will not be regarded as false positives; but nor will they count as true positives for the study endpoints, which focus on lesions of 6mm or greater as per standard CTC practice and international recommendations [16].

Both the intervention and control arms will undertake their CTC tests at baseline (timepoint zero) and then 1-, 6- and 12 months post-workshop (intervention group) or 1-, 6- and 12 months post-enrolment (control group).

### Intervention: One-day training workshop

Following the baseline assessment, the intervention arm will receive one-day, face-to-face training at a workshop delivered by our group, with invited guest faculty from across the UK to represent established sites of CTC training expertise. The training day will focus on practical teaching, using real-world, endoscopically validated CTC imaging datasets depicting a range of abnormalities. There will be no overlap between example teaching cases included in the workshop and those included in the 4 CTC tests.

We know from prior national survey data that virtually all participants will have already received basic CTC interpretation training via attendance at an introductory CTC course [14]. Therefore, we will build on this, delivering an individualised course designed to practise interpretative technique and focussing on methods to identify subtle, easily missed polyps. The workshop will follow an Objective Structured Clinical Examination (OSCE)-type format with a single CTC-related topic addressed at each station to develop an individual’s strengths and address weaknesses. Since assessment has been shown to encourage learning [17], structured one-to-one expert training by highly experienced radiographers and radiologists will be alternated with practical test cases for personal assessment.

At the workshop, each participant will receive written feedback of their baseline test performance, including details of any false positive lesions identified and anonymous benchmarking of their score against the rest of the cohort. At each topic station they will also receive verbal feedback from an Expert Trainer and review the relevant test cases to identify areas of improvement. Following each subsequent test, radiologists randomised to the intervention arm will receive their written results with feedback and will be invited to receive additional verbal feedback via a phone call (up to one hour) with a faculty member to discuss their performance (figure 1).

**Figure 1.**
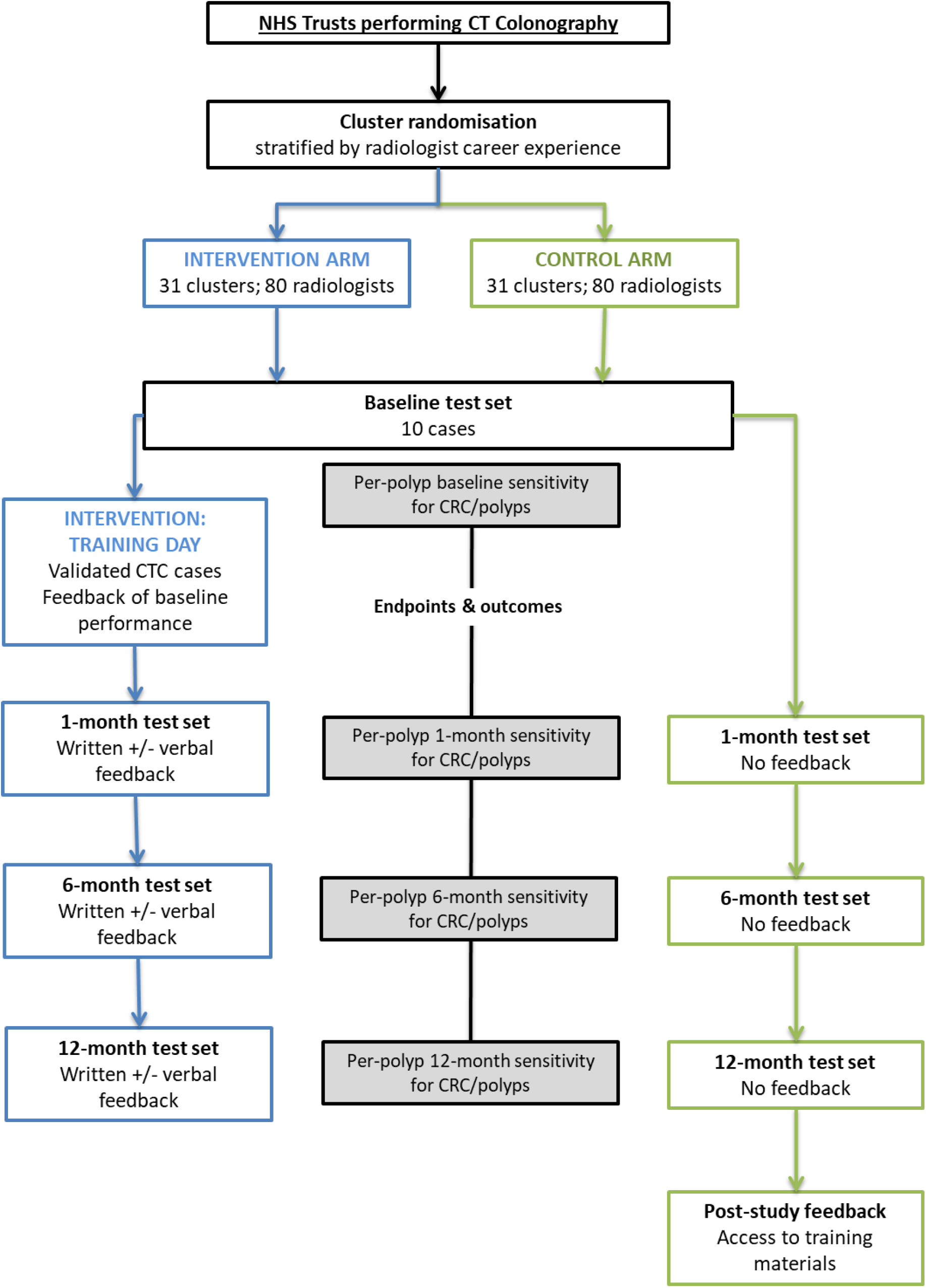
Participant timeline.

The control arm will not attend the training workshop and will not receive any written or verbal feedback of their performance. Results of their tests will only be provided at the end of the study (figure 1); however, to encourage adherence to the protocol, all participants will be able to claim CPD points for private study.

### Outcomes

While randomisation will be performed at cluster level, the intervention is targeted at individual radiologists; therefore, study outcomes will prioritize individual-level outcomes.

The primary outcome will be the difference in per-polyp sensitivity between study arms at the 1-month timepoint. Secondary outcomes include the difference in per-polyp sensitivity at 6 months and, in particular 12 months post-intervention (as 12 months represents the most realistic time point at which any repeat / refresher training would likely be administered).

Further secondary endpoints, and pre-specified subgroup analyses, are provided in the list below.

- Primary outcome: Difference in per-polyp sensitivity between intervention and controls for CRC / 6mm+ polyps at the 1-month post-training test set.
- Secondary outcomes:
  ○ Difference in per-polyp sensitivity between intervention and controls for CRC / polyps at the 6- and 12-months post-training test set (to test longer-term durability of one-off training)
  ○ Difference in per-case sensitivity pre- and post-intervention
  ○ Difference in per-case specificity pre- and post-intervention
  ○ Number of false positives per case pre- and post-intervention
  ○ Radiologist characteristics associated with higher sensitivity; including career experience and use of 3D reporting
  ○ Polyp characteristics associated with higher detectability by radiologists; including morphology, segmental location and size
  ○ Characteristics of false positive diagnoses; including morphology, segmental location and size.

We will also examine baseline radiologist characteristics between the two arms and individual clusters, as well as differences in per-polyp and per-case sensitivity at baseline between the two trial arms, to ensure the randomised groups are acceptably well-matched.

### Sample size

A survey of current practice in the BCSP identified an average of 3 radiologists per hospital, with a total of 400 radiologists in 110 hospitals [18]. We anticipate a median cluster size of 2, as we do not expect that all radiologists in a given centre will have time to participate in the study.

Our sample size calculation is based on a comparison of the sensitivity between the two study arms. A sensitivity of 70% is assumed in the control arm, and we regard a 10% increase (i.e. to 80%) in the intervention arm as being of clinical importance. If sensitivity for a given polyp were assumed to be independent data points, then with a 5% significance level and 80% power, the study would require 294 events (i.e. polyps or cancers) per arm. However, as there are multiple cases per radiologist and multiple radiologists per centre, this requires that the sample size is increased to account for the non-independence of these data. This inflation factor (i.e. the design effect) is 1 + ρ(n-1) [19], where ρ = Intracluster Correlation Coefficient (ICC) and n = number of positive cases interpreted by each radiologist. Previous data from CTC reader performance studies using computer-aided detection suggest an ICC of 0.092 [20, 21]. Therefore, using this ICC value, and the number of positive cases, n=6, the design effect for this study will be 1.46.

Accordingly, we require 429 positive CTC cases per arm (i.e. 294*1.46). We intend to include 10 CTC examinations per radiologist, at a prevalence of 60%; this implies the total number of cases to be read will be 715 (i.e. 429/0.6). Since each radiologist will read 10 cases, this requires a sample size of 72 radiologists per arm. To allow for a 10% drop-out, it is intended to recruit 80 radiologists per arm into the study (160 in total).

As there will be multiple polyps for some cases, a comparison of the per-polyp sensitivity between groups is likely to have >80% power to detect a similar-sized difference of 10% between treatment groups.

### Recruitment

The study will be advertised at the British Society of Gastrointestinal Radiologists (BSGAR) annual conference and on its own specifically-designed website [22]. We anticipate that most exposure will come from word of mouth, and we will encourage recruited participants to invite their colleagues to take part.

### Assignment of intervention

To ensure balanced baseline characteristics between the control and intervention arms, we will use stratified randomisation, according to career experience of more or less than 1000 CTCs reported, as per the first radiologist recruited for each cluster. Radiologist characteristics, including career experience, mode of reporting (2D vs 3D), BCSP reader and time spent reporting will be collected via online questionnaire prior to randomisation.

Within each stratum, pseudo-random numbers will be used to allocate clusters to each of the two study groups. Blocked randomisation, with varying block sizes, will be used to ensure a balance of clusters in each group throughout the randomisation process.

The research co-ordinator will enrol the radiologists and administer the pre-randomisation questionnaire. Radiologist career experience will be collated and forwarded to the trial statistician who will generate the allocation sequence. Radiologists assigned to the intervention arm will be contacted by the research co-ordinator and invited for the one-day workshop following completion of their baseline test.

### Data collection and management

The pre-randomisation questionnaire and test CRF will be distributed via an online, password protected survey tool. Each radiologist will retain an anonymous participant ID for the duration of the study, and anonymous data will be exported into Microsoft Excel spreadsheets and tests marked manually by the central research team. The original participant CRFs will be downloaded and archived as study source data.

Participants will be given a two- to three-week window within which to complete each CTC test and retention will be encouraged by automatically generated reminders sent from the survey tool. Outcome data will be collected for every completed test, even if a participant discontinues the study before the final assessment.

### Statistical Methods

The primary outcome is the per-polyp sensitivity at one-month. To allow for the data structure, the treatment groups will be compared using multilevel logistic regression. As each radiologist assesses the same cases/polyps, a cross-classified model will be used. Individual polyps will be nested within cases, which will be cross-classified with radiologists. The radiologists will be nested within centres. The per-polyp sensitivity of the radiologist at baseline will be included as a covariate in the model. The analysis will be restricted to segments where a polyp/CRC meeting the defined criteria is observed. Similar methods will be utilised to compare per-polyp sensitivity between groups at 6 and 12-months post-training.

A similar approach will be used to analyse the per-case sensitivity. Multilevel logistic regression will again be utilised, using a simpler model with only one measurement per case (i.e. detection or not of the index lesion, defined as per above). Only cases with polyps/CRC meeting the criteria will be included in the analysis.

The number of false positives per patients will also be analysed using multilevel statistical methods. To allow for the likely heavily positively skewed distribution, a Poisson or negative binomial model will be used.

Exploratory analyses examining factors associated with the main study outcomes will use equivalent methods to those for the group comparisons.

Missing data is expected to be minimal; if required, we will use multiple imputation under the missing at random (MAR) assumption using a chained equation method and a minimum of 10 imputations.

## DISCUSSION

CTC is a widely available and commonly-used diagnostic test, with comparable diagnostic accuracy to colonoscopy in large randomised research trials; however, this is poorly replicated in clinical practice. Currently, there are no restrictions on who can report CTC and no consensus performance indicators. Lack of accreditation and established minimum reporting standards may contribute to variability in detection rates among radiologists. This study will establish whether a structured one-day training programme targeted at radiologists who already report CTC as part of their routine practice can improve their polyp detection rate. Through a series of CTC test cases, focussed on subtle/easily missed lesions we will determine whether assessment, training and feedback can have a positive impact versus just assessment alone.

While there is consensus that CTC accreditation would be beneficial [14], the exact format of this must be determined. This study will be of value in proposing a potential model for accreditation (involving testing and training), with associated guidance on key performance indicators and factors associated with improved radiologist accuracy.

## Data Availability

The datasets during and/or analysed during the current study will not be made publicly available.

## ETHICS AND DISSEMINATION

### Ethical approval

Local ethical approval has been obtained from the University College London Research Ethics Committee (5967/003; 21 June 2016) and approval from the Health Research Authority (HRA) was obtained on 2 Dec 2016 (IRAS ID: 206876).

### Consent

Participant consent will be obtained in writing by the research co-ordinator following review of a participant information sheet by each eligible radiologist.

### Availability of data and materials

The datasets during and/or analysed during the current study will not be made publicly available.

### Competing interests

AAP: No competing interests

AO: No competing interests

PB: No competing interests

RB-C: No competing interests

SH: Provides non-remunerated research and development advice for iCAD inc.

DB: No competing interests

### Dissemination policy

Individual participant results will be shared with the intervention arm during the study as per the protocol and individual control arm results will be shared following study completion. Comparison of the results and full statistical analysis will be shared via publication.

## Authors’ contributions

AAP conceived the study and wrote the protocol with AEO, DNB, RB-C and SH. PB designed the statistical analysis plan. AEO and AAP drafted the manuscript. All co-authors edited, revised, and contributed to the intellectual content of the manuscript. The views presented in this protocol are those of the authors, and not necessarily those of the NIHR, Department of Health, St Mark’s Hospital Foundation, 40tude, the Edith Murphy Foundation or Public Health England.

